# Internalizing Problems Before and During the COVID-19 Pandemic in Dutch Children and Adolescents with and without Pre-Existing Mental Health Problems

**DOI:** 10.1101/2021.10.05.21264160

**Authors:** Karen Fischer, Jacintha M. Tieskens, Michiel A. J. Luijten, Josjan Zijlmans, Hedy A. van Oers, Rowdy de Groot, Daniël van der Doelen, Hanneke van Ewijk, Helen Klip, Rikkert M. van der Lans, Ronald De Meyer, Malindi van der Mheen, Maud M. van Muilekom, I. Hyun Ruisch, Lorynn Teela, Germie van den Berg, Hilgo Bruining, Rachel van der Rijken, Jan Buitelaar, Pieter J. Hoekstra, Ramón Lindauer, Kim J. Oostrom, Wouter Staal, Robert Vermeiren, Ronald Cornet, Lotte Haverman, Meike Bartels, Tinca J. C. Polderman, Arne Popma

## Abstract

The aim of the study was to assess internalizing problems before and during the pandemic with data from Dutch consortium *Child and adolescent mental health and wellbeing in times of the COVID-19 pandemic*, consisting of two Dutch general population samples (GS) and two clinical samples (CS) referred to youth/psychiatric care. In each sample, measures of internalizing problems were obtained from ongoing data collections pre-pandemic (*N*_GS_= 35,357; *N*_CS_= 4,487) and twice during the pandemic, in Apr.–May 2020 (*N*_GS_= 3,938; clinical: *N*_CS_= 1,008) and in Nov.–Dec. 2020 (*N*_GS_= 1,489; *N*_CS_= 1,536), in children and adolescents (8-18 years) with parent- (Brief Problem Monitor) and/or child reports (Patient-Reported Outcomes Measurement Information System^®^). Results show significantly greater proportions of worrisome internalizing problems (based on validated cut-offs) and significantly higher internalizing problems mean levels from pre-pandemic to pandemic measurements in the general population. These levels stabilized or decreased over the course of the pandemic. In the clinical sample, we found an increase in child-reported internalizing problems measures over the course of the pandemic, but parents reported no differences in internalizing problems measures over the course of the pandemic, nor from pre-pandemic to during the pandemic. Overall, the findings indicate that children and adolescents of both the general and clinical population were affected negatively by the pandemic in terms of their internalizing problems. Attention is therefore warranted to investigate what long-term effects this may cause and to monitor if internalizing problems return back to pre-pandemic levels or if they remain elevated post-pandemic.

---

The implemented social distancing measures during the COVID-19 pandemic have brought about marked changes in the daily lives of individuals and families across the globe. Restrictions such as primarily working at home, closure of schools and social clubs, and limited physical contact with friends and even family members have characterized life during the COVID-19 pandemic lockdown. Reviews synthesizing results of longitudinal and cross-sectional studies on adult’s mental health during the COVID-19 pandemic illustrate a general picture of psychological resilience and little decline in mental health in the general population [1]. However, for certain populations, such as females, or those with pre-existing mental health problems, studies indicate increased –albeit not clinical– levels of stress, depression, and anxiety since the onset of lockdown measures [2]. The effects of the restrictions are especially of concern regarding the psychosocial development of children and adolescents, since social interactions and forming relationships with peers –which were both limited during the COVID-19 pandemic– are crucial components of a healthy development during this age [3]. Social deprivation may contribute to feelings of loneliness, disconnection from one’s peers, and experiencing depressive and anxious feelings [4]. In addition, the fear of the virus itself and the uncertainty of how this might affect one’s family or the world in general may also negatively affect children’s and adolescents’ mental health [5].

Several cross-sectional studies from China conducted in children and adolescents in the general population [6–8] indicated higher prevalence of anxiety and depressive symptoms during the first lockdown than pre-pandemic, however these differences were not statistically assessed. Initial results from one of our general population-based samples [9] are in line with these findings, showing that children and adolescents (N= 844) reported more anxiety and depressive symptoms during the first COVID-19 lockdown in the Netherlands (Apr. 2020), compared to a reference sample before the pandemic. Similarly, another population-based study in Germany (N= 1556), also using a reference sample as a pre-pandemic measure, found that two-thirds of children reported significantly more mental health problems and a decline in health-related quality of life since lockdown began [10]. Longitudinal studies up to this date corroborate this pattern. For example, a study from the UK (N= 168) showed that children (aged 7.6–11.6) reported an increase in depressive symptoms during the first lockdown, when compared to their ratings 18 months earlier before the pandemic, and that this effect did not significantly differ across age, gender, and family SES [11]. Or for example, a longitudinal study in 248 adolescents showed that self-reported depressive and anxiety symptoms were significantly higher two months into the pandemic than in the year preceding the pandemic [12]. Another longitudinal study with data of 12 longitudinal studies from the US, The Netherlands and Peru compiled (N= 1,339), showed an increase in depressive symptoms from pre-pandemic to the first half year of the pandemic in children and adolescents aged 9-18 years old [13].

As these studies were conducted exclusively in the general population, it remains less clear how the pandemic affects children’s internalizing problems in vulnerable groups, such as those with pre-existing mental health problems. Initial findings from our group [14] showed that during the pandemic children in psychiatric care self-reported more depressive symptoms, but not more anxiety than children from the general population. A recent systematic review on the effects of the pandemic on adolescent mental health shows that adolescents with pre-existing mental health conditions experienced a worsening in their pre-existing conditions with onset of the pandemic [15].

In light of this literature, studies using larger and more diverse samples –ranging from general to referred clinical populations– are necessary in order to yield a clearer picture regarding variations and divergence in mental health in children and adolescents before and during the pandemic. To gain such insights, we investigated the effects of the COVID-19 pandemic on internalizing problems in children and adolescents between 8-18 years with and without pre-existing mental health problems, in four individual samples: two large Dutch population-based and two clinical samples. Specifically, we assessed child- and parent-reported internalizing problems before and over the course of the pandemic in these samples with 1) mean level changes in internalizing problems and 2) proportions of worrisome internalizing problems, to determine whether more children require additional (clinical) support. Respective insights may provide important information for policy makers and mental health prevention- and intervention services in times of the COVID-19-or potential future pandemics.

## Methods

### Participants

The present study used data from children and adolescents of 8-18 years from the Dutch consortium *Child and adolescent mental health and wellbeing in times of the COVID-19 pandemic (CAMHWB-19)*, which is a unique Dutch collaboration consisting of four large child and adolescent samples: two general population-based ongoing cohort studies (*N*_pre-pandemic_= 35,357; *N*_Apr.-May 2020_= 3,938; *N*_Nov-Dec 2020_= 1,489), namely the Netherlands Twin Register (**NTR**, [16]), and a representative sample of the Dutch population (**KLIK)**, and two clinical populations (*N*_pre-pandemic_= 4,487; *N*_Apr.-May 2020_= 1,008; *N*_Nov-Dec 2020_= 1,536), namely Dutch REsearch in child and Adolescent Mental health (**DREAMS)**, which consists of children and adolescents receiving mental health care in four academic child and adolescent psychiatry centers (Amsterdam, Groningen, Leiden, Nijmegen), and **Learning Database Youth** (**LDY**), which is a collaboration between youth care centers in the Netherlands that collects data about mental health status of patients during their treatment trajectory. Table 1 gives an overview of the sample sizes, respective ratios of boys versus girls, type of rater (mother or father), and average age of participants per sample. An extensive description of the individual samples and respective details of data collection procedures can be found in the Supplementary Materials.

**Table 1.** Sample size and respective percentage of boys, mean age and standard deviations before and during the pandemic for BPM and PROMIS in all samples.

| Sample | Pre-pandemic |  |  |  |  | Pandemic<br>Apr.–May 2020 |  |  |  |  | Pandemic<br>Nov.–Dec. 2020 |  |  |  |  |
| --- | --- | --- | --- | --- | --- | --- | --- | --- | --- | --- | --- | --- | --- | --- | --- |
| | N | %<br>males | %<br>mother<br>ratings | $M_{age}$ | $SD_{age}$ | N | %<br>males | %<br>mother<br>ratings | $M_{age}$ | $SD_{age}$ | N | %<br>males | %<br>mother<br>ratings | $M_{age}$ | $SD_{age}$ |
| **General Population Samples** |  |  |  |  |  |  |  |  |  |  |  |  |  |  |  |
| *Brief Problem Monitor (parent-reported internalizing problems)* |  |  |  |  |  |  |  |  |  |  |  |  |  |  |  |
| NTR | 34038 | 49.5% | 97.7% | 10.4 | (1.9) | 3524 | 53.7% | 84.7% | 9.2 | (2.1) | 1168 | 49.0% | 88.1% | 9.8 | (2.3) |
| *PROMIS (child-reported internalizing problems)* |  |  |  |  |  |  |  |  |  |  |  |  |  |  |  |
| KLIK | 1319 | 49.4% | N/A* | 12.7 | (3.1) | 832 | 46.3% | N/A* | 13.4 | (2.8) | 746 | 53.3% | N/A* | 13.7 | (3.2) |
| **Clinical Samples** |  |  |  |  |  |  |  |  |  |  |  |  |  |  |  |
| *Brief Problem Monitor (parent-reported internalizing problems)* |  |  |  |  |  |  |  |  |  |  |  |  |  |  |  |
| DREAMS | 1395 | 61.9% | N/A** | 11.4 | (2.4) | 453 | 55.6% | 79.9% | 13.1 | (3.1) | 726 | 58.5% | 84.4% | 13.2 | (2.9) |
| LDY | 3092 | 62.3% | 68.8% | 13.2 | (3.0) | 280 | 62.5% | 61.4 % | 13.4 | (2.9) | 302 | 64.6% | 53.0 % | 13.7 | (3.0) |
| *PROMIS (child-reported internalizing problems)* |  |  |  |  |  |  |  |  |  |  |  |  |  |  |  |
| DREAMS | - | - | - | - | - | 275 | 54.2% | N/A* | 13.0 | (3.1) | 508 | 50.2% | N/A* | 13.9 | (2.9) |

Note. N= Sample size, % males= percentage of males in sample, % mother ratings= percentage of mother ratings in sample, $M_{age}$ = mean age, $SD_{age}$ = standard deviation age. BPM= Brief Problem Monitor, PROMIS= Patient-Reported Outcomes Measurement Information System. \*not applicable due to child report measure; \*\*not applicable due to unknown informant.

### Design and Procedure

The present study employed a between-subject design, with data before and during the pandemic from four collaborating parties (NTR, KLIK, DREAMS, LDY). Each party collected cross-sectional data assessing child- and/or parent-reported internalizing problems in independent samples pre-pandemic and twice during the pandemic (with the exception of DREAMS which does not have a child-reported pre-pandemic measurement). If a participant had participated on more than one of the three measurements, inclusion of only one of the respective measurements was determined with randomization, to maintain independent samples. Measures of pre-pandemic internalizing problems were obtained from the ongoing data collections that took place at various time points before the pandemic within the individual samples (collected anywhere between 2018 and 2019, with the exception that for NTR some of the pre-pandemic assessments reached back as far as 1995). For the first pandemic measurement, all collaborating parties collected data in the same time window of Apr.–May 2020, the first peak of the pandemic during which there was a strict lockdown in the Netherlands. Some of the main social distancing measures in Apr.–May in the Netherlands included working from home, closure of schools, sports- and social clubs. For the second pandemic measurement, all collaborating parties collected data in the same time window between Nov. – Dec. 2020, during which there was a partial lockdown (when schools had reopened) in the Netherlands. Figure 1 presents a timeline of the most important regulations that were active at the time of the measurement points during the pandemic. Prior to the start of the study, collaborating parties received approval for data collection by the appropriate ethics committees, and all children and parents provided informed consent. Data from the LDY sample were not collected specifically for the current study but as part of patients’ treatment trajectory. The studies were conducted in line with the ethical standards stated in the 1964 Declaration of Helsinki and its later amendments.

**Fig 1.**
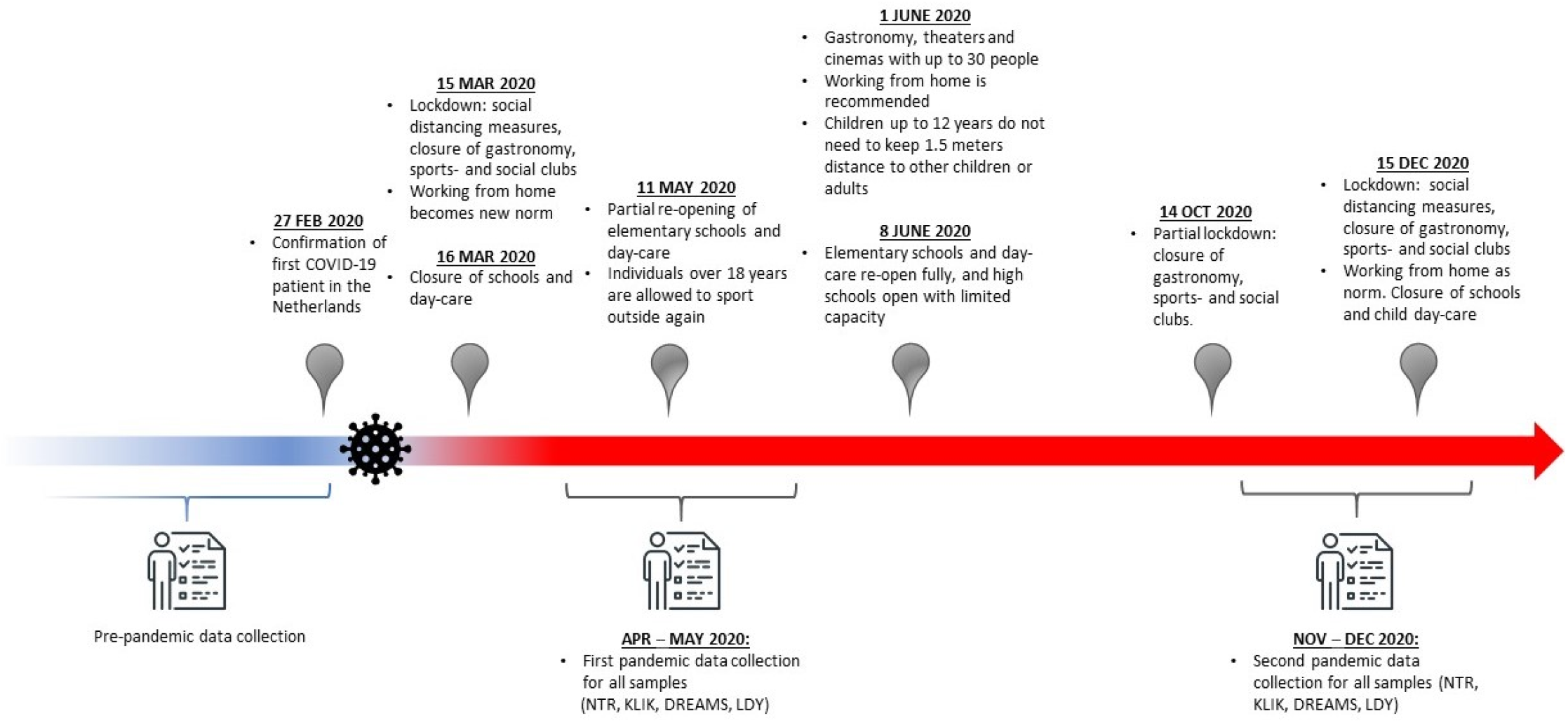
Timeline COVID-19 Regulations in the Netherlands.

### Measures

#### Parent-Reported Internalizing Problems

##### Brief Problem Monitor (BPM)

The BPM [17] is a shortened version of the Child Behavior Checklist (CBCL/6-18 years; [18]), which is a parent report questionnaire measure of behavioral and emotional problems in children. To assess parent-reported internalizing problems, the internalizing problems scale of the BPM [17] was used, consisting of 6 items. Items were rated on a three-point Likert-scale, asking parents to rate how much a statement applies to their child (0 = ‘not true’, 1 = ‘somewhat true’, to 2 = ‘very true’). The BPM contains five items assessing anxious/depressed symptoms (e.g., “worries”), and one item assessing withdrawn/depressed internalizing problems (e.g., “unhappy, sad, or depressed”). Items were summed to yield a scale score, where higher scores signify more internalizing problems. In line with the BPM manual, missing items on the BPM internalizing problems scale were coded as zero [17]. If more than 20% of items were missing on the BPM internalizing problems scale, the scale was set as missing for that individual. This applied to only NTR and LDY data, as the samples KLIK and DREAMS did not have missing data, as all questions were mandatory.

#### Child-Reported Internalizing Problems

##### Patient-Reported Outcomes Measurement Information System (PROMIS^®^)

The Dutch-Flemish PROMIS^®^ (Patient-Reported Outcomes Measurement Information System) pediatric V2.0. Item Bank Anxiety and V2.0. Item Bank Depressive Symptoms were used to assess child-reported internalizing problems and are developed using modern psychometric techniques [19] that measure their respective domains of anxiety and depressive symptoms in children. The Anxiety and Depressive Symptoms [20] item banks were administered as Computerized Adaptive Tests (CAT), where items are selected based on responses to previously completed items, resulting in a reliable score with a few items. The anxiety item bank contains 15 items that reflect fear (e.g., fearfulness), anxious misery (e.g., worry), and hyperarousal (e.g., nervousness) [20]. The depressive symptoms item bank contains 14 items on negative mood (e.g., sadness), anhedonia (e.g., loss of interest), negative views of the self (e.g., worthlessness, low self-esteem), and negative social cognition (e.g., loneliness, interpersonal alienation) [20]. All PROMIS measures use a 7-day recall period, and most items are scored on a five-point Likert scale ranging from ‘never’ to ‘(almost) always’. Total scores are calculated by transforming the item scores into a *T-score* ranging from 0 to 100 which has a mean of 50 and standard deviation (SD) of 10 in the U.S. calibration sample [20], where higher scores thus signify more internalizing problems. The US item parameters were used in the CAT algorithm and T-score calculations, as by PROMIS convention.

### Data Analysis

For the BPM, in accordance with the Manual for the ASEBA Brief Problem Monitor [17], T-scores of the children were interpreted as within the normal range (T<65) or as ‘elevated’ (T>=65) internalizing problems. T-scores (T= (z*10)+50) were calculated by age (6-11 years old /12-18 years old), sex (boys/girls), and rater (mother/father), based on the large-scale pre-pandemic population-based data of the NTR. Specifically, this norm sample (N = 34,038) consisted of the most recent pre-pandemic assessment of those individuals from the NTR from whom no BPM data during the pandemic were available, thereby yielding a population representative independent sample. The sex-, age- and rater-specific absolute scale score cut-offs are displayed in Table S1 in the Supplementary Materials. The sample sizes of each of the eight norm groups, and respective means and standard deviations on the BPM internalizing problems scale are displayed in Table S2 in the Supplementary Materials. We then determined which raw BPM internalizing problems scale scores corresponded to the T-score cut-offs. For PROMIS measures, we used previously defined cut-off scores based on a representative general population sample measured before the pandemic [21, 22]. The cut-off from normal to mild symptoms/function was the 75^th^ percentile and the cut-off from mild to severe was the 95^th^ percentile.

Within each sample, we performed independent t-tests to assess differences in mean levels of internalizing problems (total scores of BPM or PROMIS) between measurements (pre-pandemic, pandemic 1, pandemic 2). Then, within each sample, we performed Chi-Square *X*^2^ tests to assess differences in proportions of respondents with worrisome internalizing problems across measurements. We defined worrisome internalizing problems using the cut-offs for the BPM and PROMIS, where for PROMIS, we combined ‘mild’ and ‘severe’ symptoms into the ‘worrisome’ internalizing problems category, and for BPM the ‘elevated’ category represented the ‘worrisome’ internalizing problems category. All analyses were Bonferroni corrected for number of tests within sample, for each instrument (i.e., BPM or PROMIS). Reported p-values are uncorrected.

## Results

Table 2 displays the internalizing problem scale scores on the BPM and PROMIS anxiety and depressive symptoms before and during the pandemic in all samples. Table 3 displays the respective proportions of worrisome internalizing problem scores, as assessed with the BPM (‘elevated’ range) and PROMIS (‘mild’ and ‘severe’ range) before and during the pandemic in all samples. Although the results represent comparisons between independent, cross-sectional measurements, for simplicity differences in internalizing problems between measurements will be referred to as increases, decreases, or stabilization in internalizing behavior.

**Table 2.** Scale scores and respective standard deviations on the BPM and PROMIS Anxiety and Depressive Symptoms before and during the pandemic in all samples.

Internalizing Problems Before and During the COVID-19 Pandemic
| Sample | Pre-pandemic<br>(a) |  | Pandemic<br>Apr.–May 2020 (b) |  | Pandemic<br>Nov.–Dec. 2020 (c) |  |
| --- | --- | --- | --- | --- | --- | --- |
|  | M | SD | M | SD | M | SD |
| **General Population Samples** |  |  |  |  |  |  |
| *Brief Problem Monitor (parent-reported internalizing problems)* |  |  |  |  |  |  |
| **NTR** | .90<sup>b,c</sup> | (1.51) | 1.49<sup>a,c</sup> | (1.99) | 1.07<sup>a,b</sup> | (1.71) |
| *PROMIS Anxiety (child-reported internalizing problems)* |  |  |  |  |  |  |
| **KLIK** | 43.76<sup>b,c</sup> | (9.9) | 50.78<sup>a,c</sup> | (7.78) | 49.61<sup>a,b</sup> | (8.25) |
| *PROMIS Depressive Symptoms (child-reported internalizing problems)* |  |  |  |  |  |  |
| **KLIK** | 44.73<sup>b,c</sup> | (10.6) | 49.53<sup>a</sup> | (8.20) | 48.81<sup>a</sup> | (9.18) |
| **Clinical Samples** |  |  |  |  |  |  |
| *Brief Problem Monitor (parent-reported internalizing problems)* |  |  |  |  |  |  |
| **DREAMS** | 5.24 | (3.24) | 4.91 | (3.52) | 5.04 | (3.41) |
| **LDY** | 3.86 | (3.09) | 3.75 | (3.16) | 3.84 | (3.24) |
| *PROMIS Anxiety (child-reported internalizing problems)* |  |  |  |  |  |  |
| **DREAMS** | - | - | 51.45<sup>c</sup> | (8.98) | 54.45<sup>b</sup> | (9.21) |
| *PROMIS Depressive Symptoms (child-reported internalizing problems)* |  |  |  |  |  |  |
| **DREAMS** | - | - | 51.98<sup>c</sup> | (10.64) | 55.67<sup>b</sup> | (10.81) |

Note. <sup>a,b,c</sup>= represent significant differences ( $p < .05$ , Bonferroni corrected for number of tests for each instrument within sample) between measurements as
indicated by independent sample t-tests; *M*= mean, *SD*= standard deviation. BPM= Brief Problem Monitor, PROMIS= Patient-Reported Outcomes Measurement Information System.

**Table 3.**
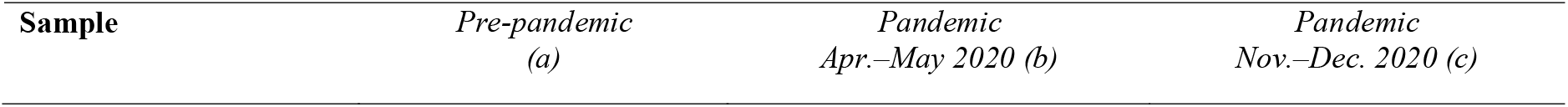
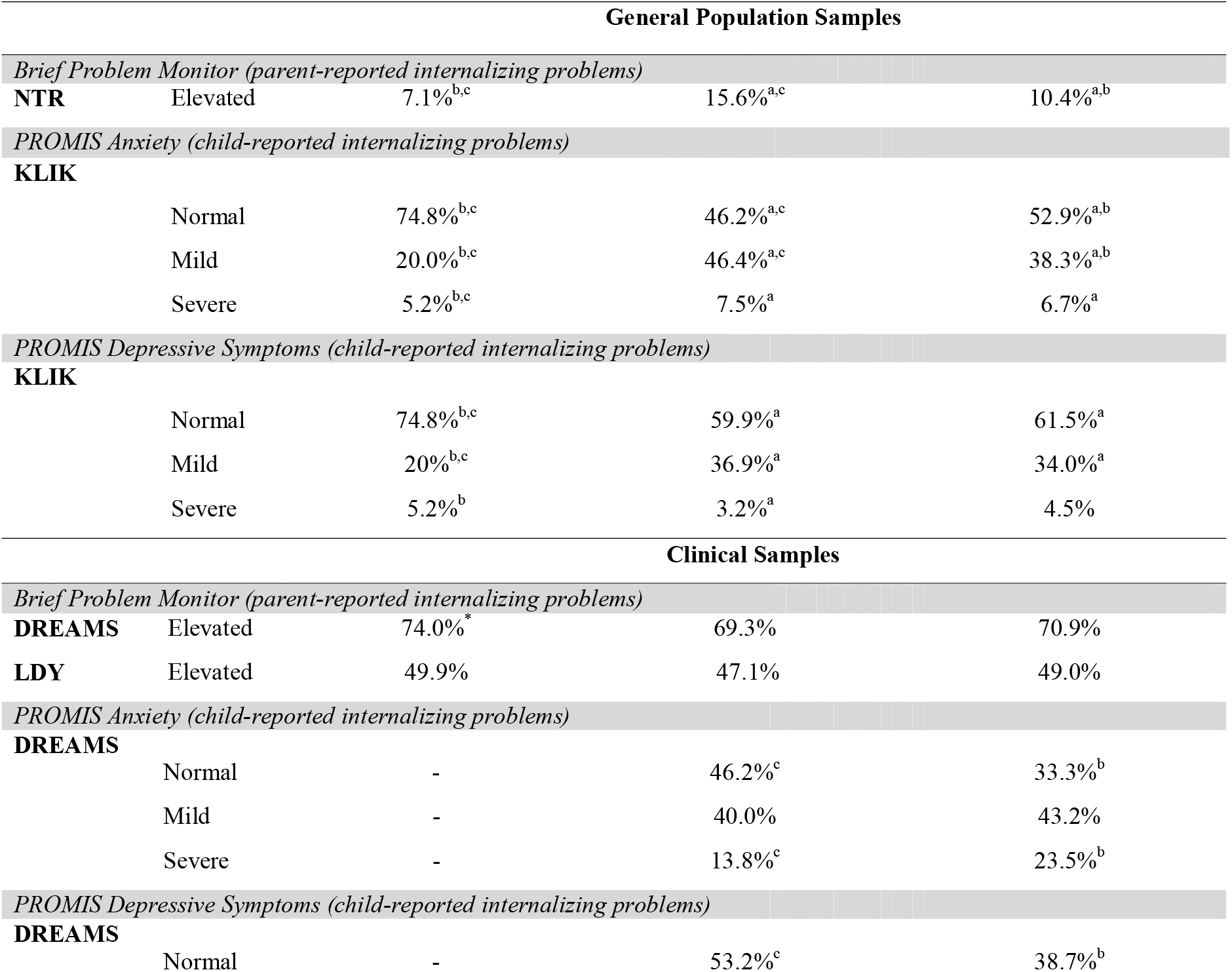
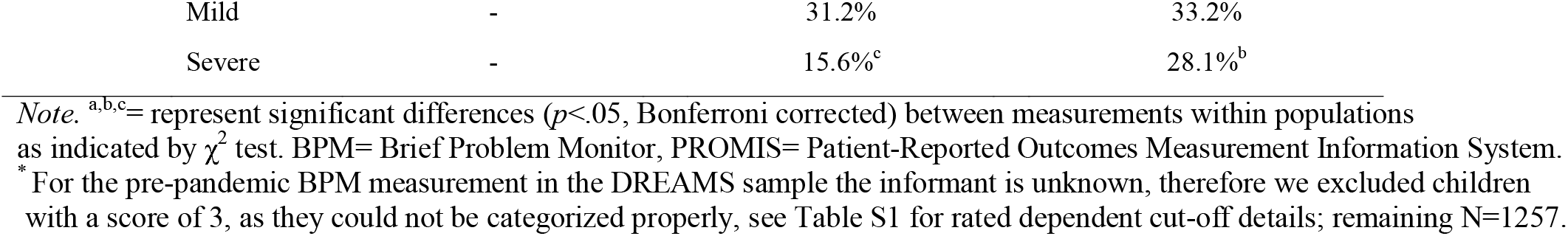
Percentages of worrisome internalizing problems as assessed with cut-off scores on the BPM and PROMIS before and during the pandemic in all samples.

| Sample | Pre-pandemic<br>(a) | Pandemic<br>Apr.–May 2020 (b) | Pandemic<br>Nov.–Dec. 2020 (c) |
| --- | --- | --- | --- |

Internalizing Problems Before and During the COVID-19 Pandemic
| General Population Samples |  |  |  |  |
| --- | --- | --- | --- | --- |
| *Brief Problem Monitor (parent-reported internalizing problems)* |  |  |  |  |
| **NTR** | Elevated | 7.1%<sup>b,c</sup> | 15.6%<sup>a,c</sup> | 10.4%<sup>a,b</sup> |
| *PROMIS Anxiety (child-reported internalizing problems)* |  |  |  |  |
| **KLIK** |  |  |  |  |
|  | Normal | 74.8%<sup>b,c</sup> | 46.2%<sup>a,c</sup> | 52.9%<sup>a,b</sup> |
|  | Mild | 20.0%<sup>b,c</sup> | 46.4%<sup>a,c</sup> | 38.3%<sup>a,b</sup> |
|  | Severe | 5.2%<sup>b,c</sup> | 7.5%<sup>a</sup> | 6.7%<sup>a</sup> |
| *PROMIS Depressive Symptoms (child-reported internalizing problems)* |  |  |  |  |
| **KLIK** |  |  |  |  |
|  | Normal | 74.8%<sup>b,c</sup> | 59.9%<sup>a</sup> | 61.5%<sup>a</sup> |
|  | Mild | 20%<sup>b,c</sup> | 36.9%<sup>a</sup> | 34.0%<sup>a</sup> |
|  | Severe | 5.2%<sup>b</sup> | 3.2%<sup>a</sup> | 4.5% |
| Clinical Samples |  |  |  |  |
| *Brief Problem Monitor (parent-reported internalizing problems)* |  |  |  |  |
| **DREAMS** | Elevated | 74.0%<sup>*</sup> | 69.3% | 70.9% |
| **LDY** | Elevated | 49.9% | 47.1% | 49.0% |
| *PROMIS Anxiety (child-reported internalizing problems)* |  |  |  |  |
| **DREAMS** |  |  |  |  |
|  | Normal | - | 46.2%<sup>c</sup> | 33.3%<sup>b</sup> |
|  | Mild | - | 40.0% | 43.2% |
|  | Severe | - | 13.8%<sup>c</sup> | 23.5%<sup>b</sup> |
| *PROMIS Depressive Symptoms (child-reported internalizing problems)* |  |  |  |  |
| **DREAMS** |  |  |  |  |
|  | Normal | - | 53.2%<sup>c</sup> | 38.7%<sup>b</sup> |
| Mild | - | 31.2% | 33.2% |
| Severe | - | 15.6%<sup>c</sup> | 28.1%<sup>b</sup> |

*Note.* <sup>a,b,c</sup>= represent significant differences ( $p < .05$ , Bonferroni corrected) between measurements within populations as indicated by $\chi^2$ test. BPM= Brief Problem Monitor, PROMIS= Patient-Reported Outcomes Measurement Information System.
\* For the pre-pandemic BPM measurement in the DREAMS sample the informant is unknown, therefore we excluded children with a score of 3, as they could not be categorized properly, see Table S1 for rated dependent cut-off details; remaining N=1257.

### General Population Samples

#### Parent Report Results

For the parent report measures on the BPM in the general population (NTR), we found that from pre-pandemic (*M* = .90, *SD* = 1.51) to during the first pandemic measurement (*M* = 1.49, *SD* = 1.99) mean levels of internalizing problems increased (*t*(37560) = -19.87, *p* < .001), see Table 2. Furthermore, we also found that from pre-pandemic to during the second pandemic measurement (*M* = 1.07, *SD* = 1.71), mean levels of internalizing problems increased (*t*(35204) = -3.61, *p* < .001). In Table 3, Chi-square tests on the proportion of internalizing scores in the ‘elevated’ range show that the group with ‘elevated’ problems increased from pre-pandemic to during the first pandemic measurement (*X*^2^ (1, *N* = 37,562) = 316.35, *p* < .001) and from pre-pandemic to during the second pandemic measurement (*X*^2^ (1, *N* = 35,206) = 18.72, *p* < .001). From the first to the second pandemic measurement, mean levels of internalizing problems decreased, *t*(4690) = 6.44, *p* < .001), and furthermore Chi-Square tests on the proportion of ‘elevated’ internalizing scores show that the group with ‘elevated’ internalizing problems decreased, *X*^2^ (1, *N* = 4,692) = 19.05, *p* < .001, see Table 2.

#### Child Report Results

For the child report measures in the general population (KLIK), we found significant increases from before (*M* = 43.76, *SD* = 9.87) to during the first pandemic measurement (*M* = 50.78, *SD* = 7.68) for anxiety (*t*(2149) = -18.45, *p* < .001). We also found significant increases from before (*M* = 44.73, *SD* = 10.62) to during the first pandemic measurement (*M* = 49.53, *SD* = 8.20) for depressive symptoms (*t*(2149) = -11.77, *p* < 0.001) (see Table 2). In Table 3, Chi-Square tests on the proportion of worrisome internalizing problem scores from before to during the pandemic show that the number of participants with mild anxiety increased significantly (pre-pandemic–first pandemic measurement *X*^2^ (1, *N* = 2,151) = 168.36, *p* < .001; pre-pandemic–second pandemic measurement: *X*^2^ (1, *N* = 2,065) = 81.87, *p* < .001) as well as the number of participants with severe anxiety (pre-pandemic–first pandemic measurement *X*^2^ (1, *N* = 2,151) = 168.36, *p* < .001; pre-pandemic–second pandemic measurement: *X*^2^ (1, *N* = 2,065) = 81.87, *p* < .001). Furthermore, the proportion of participants with mild depressive symptoms increased (pre-pandemic–first pandemic measurement *X*^2^ (1, *N* = 2,151) = 73.84, *p* < .001; pre-pandemic–second pandemic measurement: *X*^2^ (1, *N* = 2,057) = 48.79, *p* < .001).

From the first to the second pandemic measurement, anxiety decreased (*t*(1576) = 2.91, *p =* .004), but was still significantly higher (*M* = 49.61, *SD* = 8.25) than pre-pandemic level (*t*(2063) = -14.40, *p* < .001). Depressive symptoms did not change from the first to the second pandemic measurement (*t*(1576) = 1.44, *p =* .149) and remained higher (*M* = 48.81, *SD* = 9.18) than pre-pandemic levels (*t*(2063) = -9.15, *p <* .001) (see Table 2). Chi-Square tests on the proportion of worrisome scores from the first to the second pandemic measurement show that for anxiety the proportion of participants with severe symptoms remained the same (*p* > .05), the proportion of participants with mild symptoms decreased (*X*^2^ (1, *N* = 1,578) = 10.44, *p* = .001), and the proportion of participants that show no symptoms increased (*X*^2^ (1, *N* = 1,578) = 7.27, *p* = .007). For depressive symptoms, we found no differences in worrisome scores from the first to the second pandemic measurement (*p* > .05) (see Table 3).

### Clinical Samples

#### Parent Report Results

No differences were found in DREAMS and LDY on the BPM in terms of internalizing problem mean scores (*p* > .05) (see Table 2) or proportions of ‘elevated’ internalizing problems (*p* > .05) (see Table 3) between the pre-pandemic and first or second pandemic measurement, and neither between the first and second pandemic measurement.

#### Child Report Results

In the psychiatric sample (DREAMS), over the course of the pandemic we found significant increases from the first (*M* = 51.45, *SD* = 8.98) to the second (*M* = 54.45, *SD* = 9.21) pandemic measurement for anxiety (*t*(783) = 4.39, *p* < .001), and also from the first (*M* = 51.98, *SD* = 10.64) to the second (*M* = 55.67, *SD* = 10.81) pandemic measurement for depressive symptoms (*t*(768) = 4.53, *p* < .001), see Table 2. Chi-Square tests on the proportion of worrisome internalizing problem scores from the first to the second pandemic measurement showed that the group with severe anxiety (*X*^2^ = 10.48, *p* = .001) and severe depressive symptoms (*X*^2^ = 15.17, *p* < .001) increased, the group with mild symptoms remained the same (*p* > .05), and the normal group decreased (*X*^2^ = 12.54, *p* < .001 and *X*^2^ = 14.82, *p* < .001 respectively) (see Table 3).

## Discussion

In this study, we assessed internalizing problems in children and adolescents aged 8 to 18 years before the first Dutch COVID-19 pandemic lockdown, during the first peak/Dutch lockdown (Apr.–May 2020), and during the second peak/Dutch partial lockdown (Nov.–Dec. 2020) in two general population samples and two clinical samples using parent- and child reports. In the general population we found that internalizing problems increased from pre-pandemic to the first peak of the pandemic based on both child- and parent reports. Yet, over the course of the pandemic, on both child- and parent reports, we saw stabilization or even a decrease in internalizing problems. In the clinical population we saw an increase in internalizing symptoms over the course of the pandemic based on child reports while in parent reports no differences were found in internalizing symptoms from pre-pandemic to the first peak of the pandemic nor over the course of the pandemic. We found these results for mean levels of internalizing problems as well as for changes in proportions of children from normal to mild, and mild to severe levels of internalizing problems. As such, it seems a shift of the total distribution of internalizing problems occurred, rather than changes only in subgroups with particular levels of pre-existing internalizing problems.

Our findings in the general population are in line with prior research [6–8, 11–13], showing that for children in the general population internalizing problems increased from pre-pandemic to the first peak of the pandemic. At the start of the first peak of the pandemic, both children and adults experienced significant changes in their psychosocial environment due to the implementation of social distancing measures. Given that social interactions are fundamental to a healthy development in children and adolescents [3, 4], the sudden social deprivation and changes in daily routines as introduced by lockdown (e.g., closure of schools and social/sports clubs) may have contributed to the observed increase in depressive symptoms and anxiety at the start of the pandemic, as reported by both parents and children themselves. The stabilization and decrease in proportions of worrisome internalizing problems and mean levels of internalizing problems over the course of the pandemic are in line with other research that has found that anxiety and depressive symptoms subsided in adolescents of the general population in the four months after the first peak of the pandemic [23]. Specifically, concerns about home confinement and school (e.g. transitioning to online learning) have been shown to be strongly associated with increased anxiety and depressive symptoms since the onset of the pandemic [23]. In line with our findings, this suggests that relaxation of home confinement measures after the first peak of the pandemic and habituation to the new online school environment may have contributed to a decrease in children and adolescents’ anxiety and depressive symptoms over the course of the pandemic.

For the clinical population we saw an increase in child-reported mean levels of internalizing problems and proportions of worrisome internalizing problems over the course of the pandemic. Literature indicates that children in clinical populations overall have less resilience than children without pre-existing mental health problems [24]. Resilience represents the capacity to quickly adapt to adversity, and being less resilient has been associated with worse physical, mental and emotional functioning [25]. As such, children with pre-existing problems may experience more difficulties with their mental health as the pandemic continued. Furthermore, children in clinical populations may have experienced a change in treatment quality during times of the pandemic, due to increased demands on mental health services during COVID-19, which may have led again to exacerbation of internalizing problems [26]. In contrast, for parent-reported internalizing problems in the clinical sample another pattern emerged, namely that there were no differences in internalizing symptoms from pre-pandemic to the first peak of the pandemic nor over the course of the pandemic. It has been shown that parents of children with pre-existing mental health problems may be less sensitive in their perception of changes in their child’s mental health than parents of children without pre-existing problems. Specifically, literature shows that in families of child mental health patients, family routines and functioning are already substantially accommodated to the needs of the child [27, 28], whereby a stressful life change, such as the pandemic –from a parent’s perspective– may not have introduced changes significant enough to considerably alter their perception of their child’s functioning. In addition, as parent-reported internalizing problems for the clinical population were already high before the pandemic, a ceiling effect may have taken place, where increases in internalizing behavior were not captured by the BPM. Lastly, parents of children with pre-existing problems may perceive changes in their child’s mental health as less problematic, knowing that newly arising problematics will be promptly addressed within the framework of their child’s ongoing youth/psychiatric care.

This pattern of stable parent-reported internalizing problems in the clinical population diverges from clinical children’s self-reported increasing levels of worrisome internalizing problems over the course of the pandemic. Such rater-discrepancies in child-versus parent-reported internalizing problems are commonly documented throughout the literature, with meta-analytical evidence supporting overall modest agreement between child- and parent-rated internalizing behaviors for child and adolescent mental health patients [29]. For example, studies using the Child Behavior Checklist show poor to low agreement between child- and parent reports [30]. Multiple rater studies of internalizing problems throughout childhood indicate that rater-disagreement variance accounted for 35% of the individual differences in internalizing behavior. Up to 17% of this was accounted for by rater-specific views, while the remainder represents rater bias [31]. Corroborating the findings in the present study, literature shows that –across different racial and ethnic groups– parents tend to report fewer internalizing problems than their children [32, 33]. Furthermore, and in line with meta-analytical evidence, internalizing problems – in contrast to externalizing problems– may be less readily noticed by parents (or other informants), but represent a significant source of distress for children [34, 35]. This may result in greater rater discrepancies, especially in vulnerable populations, where children experience and thus report more internalizing problems than their parents [35].

Whereas in the clinical sample we saw an increase in internalizing problems as the pandemic continued, this pattern stands in contrast to the stabilization or decrease we found in the general population samples over the course of the pandemic. Specifically, given that child mental health patients may have a different psychosocial environment than children of the general population [27], the changes in government regulations throughout the pandemic (during our Nov.–Dec. pandemic measurement), such as re-opening of schools and social/sports clubs, may have favorably affected children of the general population but to a lesser extent the clinical populations. For example, more contact with peers may have contributed to fewer internalizing problems for children of the general population, whereas for children of clinical populations such peer contact may at baseline be more compromised (e.g., mental health problems may interfere with psychosocial functioning) or may not represent a correlate of improved mental health (e.g., school/peer group settings may perpetuate anxiety problems). Thus, the differences in the social environment/psychosocial functioning in these two populations may have amplified divergence in internalizing problems in these two populations over the course of the pandemic.

Some limitations of the present study need to be addressed. Firstly, there were no measurements on child reports (PROMIS) for the clinical population before the pandemic, and as such no inferences can be made of how great the initial impact of the pandemic was as experienced by children in youth/psychiatric care. Moreover, none of the samples had collected data at all measurements on both parent- and child reports, and furthermore representativeness of the samples was not checked except for the child reports in the general population (KLIK). Families participating in the NTR generally show high socioeconomic status [16], which may have resulted in a slight overestimation of differences between clinical and population samples, in line with literature showing that children and adolescents of families with higher socioeconomic status experienced fewer emotional and behavioral problems in stressful life situations [36]. However, since we compared internalizing problems at the various time points for each sample separately, not controlling for sociodemographic differences may first and foremost only have impacted generalizability. Furthermore, the mean age of children in the pre-pandemic and especially pandemic sample of the NTR is lower (childhood age range) than the mean age of the other samples (adolescent age range). In line with literature indicating that the COVID-19 pandemic may have especially perpetuated adolescents’ internalizing problems [23, 37], the NTR sample in our present study may as such have exhibited comparably smaller increases in internalizing problems before versus during the pandemic. Lastly, the samples at the various measurements in the present study are independent, whereby no inferences about longitudinal effects within subjects (changes) in internalizing behavior can be made, calling for future longitudinal research to address this.

The present study also has a number of strengths. We included large samples with children from both general and clinical population, collected both parent and child reports, and included a near-equal ratio of boys versus girls in all samples. Furthermore, data of the NTR per year since 1995 until 2018 shows that proportions of worrisome internalizing problems in the general population ranged from 5.6% to 8.8%, confirming that respective proportions as reached during the pandemic in the general population (16.6% and 13.0%) represent unusually elevated, rather than random fluctuations in proportions of internalizing problems (see Figure S1 in Supplementary Materials). In addition, we maximized the number of respondents by making use of father ratings when mother ratings were not available and taking potential rater differences into account by basing proportions of worrisome internalizing problems on rater-specific norms.

In summary, our results show that in the general population internalizing problems have significantly increased since the start of the pandemic and that significantly more children report worrisome levels of internalizing problems and may require additional support. In the clinical sample we found that child- (but not parent-) reported internalizing problems significantly increased over the course of the pandemic. Overall, the findings indicate that children and adolescents of both the general and clinical population were affected negatively by the pandemic in terms of their internalizing problems. Attention is therefore warranted to investigate what long-term effects this may cause and to monitor if internalizing problems return back to pre-pandemic levels or if they remain elevated post-pandemic. These insights, combined with future multi-informant and longitudinal research in children of both general and clinical populations, may provide relevant information for policy makers and mental health prevention- and intervention services in times of the COVID-19- or potential future pandemics.

## Supporting information

Supplemental sample descriptions and figure

## Data Availability

Data are available on reasonable request

