## Supplemental sample descriptions and figure for "Internalizing Problems Before and During the COVID-19 Pandemic in Dutch Children and Adolescents with and without Pre-Existing Mental Health Problems"

***^SUPPLEMENTARY MATERIALS^***

***General Population Samples***

**The Netherlands Twin Register (NTR).** The Netherlands Twin Register [17] was established in 1987 and collects data in twins and multiples from birth onwards. As part of the ongoing nationwide data collection of the NTR (see Appendix I for more information), mothers and fathers of 7, 10, and 12 year-old twins are invited to complete the Child Behavior Checklist (CBCL/6-18 years [18]). Given the longitudinal nature of the study, with data collection from birth onwards, and depending on the age of the twin pair (some reached age 12 while others are still younger), multiple pre-pandemic assessments at different ages are available. To prevent overlap, for every child only the most recent assessment was included. Given the longitudinal data collection at NTR, the pre-pandemic sample includes data from 1995 up to 2018, resulting in a markedly larger sample for the pre-pandemic data than for the pandemic measurements. During the pandemic (Apr.–May 2020), mothers and fathers were invited to complete the shortened version of the CBCL, the Brief Problem Monitor (BPM) [19]. To be able to include the largest available homogeneous sample, we used mother ratings, and if mother ratings were not available we used father ratings. For the NTR, the pre-pandemic measurement consisted of 34,038 children (49.5% boys), *M*_age_ = 10.4 (*SD* = 1.9), of which 97.7% were mother ratings and the rest father ratings. The first pandemic measurement (Apr.–May 2020) consisted of 3,524 children (53.7% boys), *M*_age_ = 9.2 (*SD* = 2.1), where we used mother ratings (84.7%) and father ratings if mother ratings were not available. The second pandemic measurement (Nov.–Dec. 2020) consisted of 1168 children (49.0% boys), *M*_age_ = 9.8 (*SD* = 2.3), where we used 88.1% mother ratings and the rest father ratings (see Table 1).

**KLIK.** Data were collected through a research website (www.hetklikt.nu) of the KLIK Patient-Reported Outcome Measures (PROM) portal developed specifically for this purpose (www.corona-studie.nl). Parents were asked to complete a socio-demographic questionnaire and the BPM (Nov. 2020) and children completed the PROMIS questionnaires (Apr. and Nov. 2020). The samples are representative of the Dutch general population within 2.5% on most key demographics (age, sex, ethnicity, region, and educational level) compared to population numbers in 2017 [20]. PROMIS data collected during the COVID-19 pandemic [10] were compared to a reference sample of 1319 children (49.4% boys), *M*_age_ = 12.7 (*SD* = 3.1) collected pre-pandemic in 2018 [20]. Detailed data collection procedures for all measurements are described in these articles. During the pandemic, PROMIS date were collected in April 2020, in a sample of 832 children (46.3% boys), *M*_age_ = 13.4 (SD = 2.8), and in November 2020, in a separate sample of 746 children (53.3% boys), *M*_age_ = 13.7 (SD = 3.2) (see Table 1).
***Clinical Samples***

**DREAMS (Dutch REsearch in child and Adolescent Mental health).** DREAMS is a collaboration between four academic child and adolescent psychiatric centers in the Netherlands (Amsterdam, Groningen, Leiden, Nijmegen) together covering the northern, western, and eastern part of the Netherlands. Participants (children receiving psychiatric care and their parents) were invited to participate by e-mail through the psychiatric center where they receive care. As with the KLIK sample, data were collected through a research website. Parents were asked to complete a socio-demographic questionnaire and the BPM, and children completed the PROMIS questionnaires. DREAMS administered the BPM and the PROMIS twice during the pandemic, (see Table 1). For the BPM, the pre-pandemic measurement consisted of 1395 children (61.9% boys), *M*_age_ = 11.42 (*SD* = 2.41), informant unknown. The first pandemic measurement (Apr.–May 2020) consisted of 453 children (55.6% boys), *M*_age_ = 13.1 (*SD* = 3.1), 79.9% mother ratings, 15.7% father ratings and 4.4% others. The second pandemic measurement (Nov.–Dec. 2020) consisted of 726 children (58.5% boys), *M*_age_ = 13.2 (*SD* = 2.9), 84.4% mother ratings, 11.6% father ratings and 4.0% others. For PROMIS, the first pandemic measurement (Apr.–May 2020) consisted of 275 children (54.2% boys), *M*_age_ = 13.0 (*SD* = 3.1). The second pandemic measurement (Nov.–Dec. 2020) consisted of 508 children (50.2% boys), *M*_age_ = 13.9 (*SD* = 2.9) (see Table 1).

**Learning Database Youth (LDY).** Learning Database Youth (LDY) is a cooperation between youth care centers in the Netherlands to collect data on the mental health status of children and adolescents who receive youth care, to improve quality of care. In this study, data of 14 youth care institutions were used. The youth care centers are situated in northern (12.7%), eastern (60.8%), southern (2.2%) and western (24.3%) parts of the Netherlands. Participating children and adolescents in the LDY sample receive youth care for various problems such as mental, pedagogical, or educational problems. Data collection was part of their treatment trajectory, where caregivers were asked to fill out questionnaires before, during, and at the end of treatment. The BPM was administered before and during the pandemic by the caregiver(s) of the child (see Table 1). To be able to include the largest available homogeneous sample, we used mother ratings. If mother ratings were not available, we used father ratings. If both mother- and father ratings were not available we used the report of another caregiver, including grandparents, adoption parents, stepparents, and foster parents. The pre-pandemic sample (Jan-Dec 2019) consisted of 3,092 children (62.3% boys), *M*_age_ = 13.2 (*SD* = 3.0), 68.8% mother ratings, 12.5 % father ratings and 18.7% others. The first pandemic measurement (Apr.–May 2020) consisted of 280 children (62.5% boys), *M*_age_ = 13.4 (*SD* = 2.9), 61.4 % mother ratings, 14.3 % father ratings and 24.3 % others. The second pandemic measurement (Nov.–Dec. 2020) consisted of 302 children (64.6% boys), *M*_age_ = 13.7 (*SD* = 3.0), 53.0 % mother ratings, 12.3 % father ratings and 34.7 % others.

**Table S1.** *Absolute scale score cut-offs on the internalizing problems scale of the BPM corresponding
to ‘normal’ (T<65)* *and ‘elevated’ (T>=65)* *internalizing problems based on NTR norm sample.*

| **Cut-off** |  | | | | | | | | | |
| --- | --- | --- | --- | --- | --- | --- | --- | --- | --- | --- |
|  | **Boys** | |  |  | | **Girls** | | | | |
|  | 6-11 years old | 12-18 years old |  | |  | | 6-11 years old | | | 12-18 years old |
| **Mother-rated** |  |  |  | |  | | | |  | |
| Normal | < 4 | < 4 | | < 4 | | | | < 4 | | |
| Elevated | >= 4 | >= 4 | | >= 4 | | | | >= 4 | | |
| **Father-rated** |  |  | |  | | | |  | | |
| Normal | < 3 | < 3 | | < 3 | | | | < 3 | | |
| Elevated | >= 3 | >= 3 | | >= 3 | | | | >= 3 | | |

Note: For the pre-pandemic BPM measurement in the DREAMS sample the informant is unknown, therefore we excluded children with a score of 3, as they could not be categorized properly.

**Table S2.** *Sample sizes of each of the eight norm groups, and respective means and standard deviations on the BPM internalizing problems scale.*

|  | **Mother-rated** | | |  | **Father-rated** | | | |
| --- | --- | --- | --- | --- | --- | --- | --- | --- |
|  | *N* | *M* | *SD* |  | *N* | *M* | *SD* | |
| **Boys** |  |  |  |  | |  |  | |
| 6-11 years old | 9,640 | .90 | 1.50 |  | 6,489 | .63 | | 1.22 |
| 12-18 years old | 6,691 | .83 | 1.46 |  | 5,296 | .64 | 1.30 | |
| **Girls** |  |  |  |  |  |  |  | |
| 6-11 years old | 9,604 | .95 | 1.53 |  | 6,433 | .63 | 1.20 | |
| 12-18 years old | 6,824 | .94 | 1.56 |  | 5,431 | .64 | 1.21 | |

*Note. N*=Sample size, *M*=mean, *SD*= standard deviation.

**Fig. 1S** Data of the NTR on the BPM per year since 1995 until 2018, and pre-pandemic (latest assessments before pandemic), first pandemic measurement (Apr.–May 2020), and second pandemic measurement (Nov–Dec. 2020) of proportions of normal and worrisome internalizing problems (left), and only worrisome problems (scaled-larger, right)

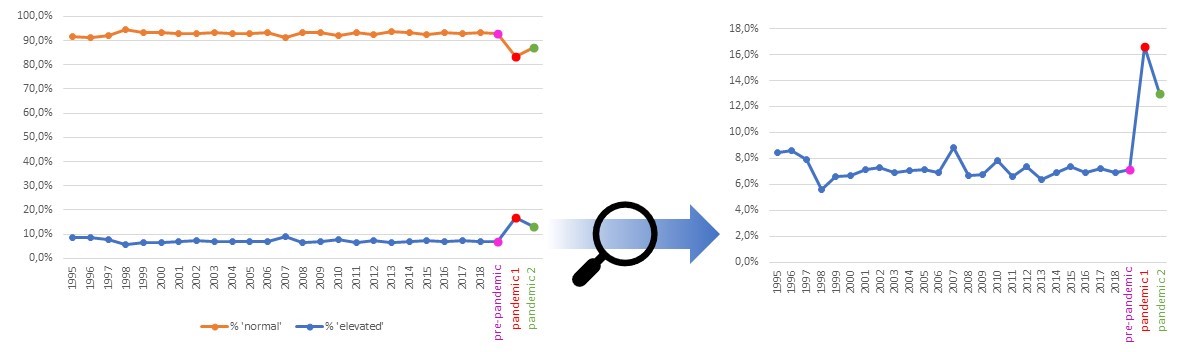
